# A Scalable Neuroinformatics Pipeline for Harmonizing Routine Clinical EEG Across Public Hospitals

**DOI:** 10.64898/2026.07.03.26357250

**Authors:** Vasily A. Vakorin, Alexander Moiseev, Sam M. Doesburg, Pengcheng Xi, Joel S. Winston, Mark P. Richardson, Roman Rodionov, Sylvain Moreno, Urs Ribary, George Medvedev

## Abstract

We propose a study protocol for routine clinical electroencephalograms (EEGs) from public hospitals, which represents a vast resource for neuroscience research. These non-invasive measures of brain function, paired with rich clinical annotations from large and diverse patient populations, are critical for developing robust artificial intelligence (AI) models and conducting population-level studies. This protocol presents a scalable methodology for curating and harmonizing extensive clinical EEG datasets, encompassing over 40,000 individual studies, to facilitate research applications. Key steps include: (i) integration of raw EEG recordings with corresponding clinical records, including neurological reports, diagnostic codes, and potentially medication data; and (ii) spatial standardization of EEG signals by mapping them to a common brain space defined by functional and anatomical landmarks. The resulting harmonized datasets enable the development of large-scale EEG foundation models, the discovery of novel EEG waveform representations, and the creation of normative “brain charts” for electrophysiological assessment across the lifespan. By enabling standardised, large-scale analyses of real-world clinical EEG data, this protocol supports data-intensive solutions for EEG applications and addresses the challenge of generalising AI models. Our approach promotes the translation of AI tools from research to diverse patient populations, advancing population neuroscience.

## Introduction

### Background: Routine Clinical EEG in Population Neuroscience

Electroencephalography (EEG), a non-invasive technique renowned for its high temporal resolution, offers direct, real-time insights into brain function and is integral to clinical practice (Michel and Murray, 2012). It plays a crucial diagnostic role in neurology and psychiatry, aiding in the assessment of conditions like epilepsy, sleep disorders, and altered consciousness (Niedermeyer, 2003). Within this clinical framework, routine EEGs stand out. They are conducted using standardized protocols, interpreted by trained neurologists, and consistently paired with detailed clinical information such as diagnoses and patient outcomes (Tatum, 2022). This systematic collection across diverse patient populations in numerous hospitals makes routine clinical EEG an unparalleled data source for understanding brain activity at scale. The resulting large-scale datasets offer a unique opportunity to advance population neuroscience, enabling studies of brain function and dysfunction across the lifespan and diverse clinical cohorts (Tveit *et al*., 2023; Beniczky *et al*., 2025; Sun *et al*., 2025). To fully harness this opportunity, robust protocols for data preparation and harmonization are essential.

EEG generates highly sampled time series data from multiple sensors placed on the scalp. These sensors capture electrical activity from various brain regions, often spanning across multiple cortical and subcortical areas (Nunez and Srinivasan, 2006). In certain applications, virtual sensors are used to reconstruct neuronal activity in specific regions of interest, providing a more focused analysis of brain dynamics (Michel and Brunet, 2019). Neuronal activity occurs across multiple brain regions simultaneously, and these regions are interconnected in dynamic networks (Buzsáki and Watson, 2012). The activity measured by EEG is not confined to a single brain area, but rather reflects interactions between local and distant regions. Also, EEG signals span multiple time scales, ranging from fast, momentary spikes to slower, rhythmic oscillations, creating a rich temporal structure (Buzsáki and Mizuseki, 2014). These characteristics make EEG signals highly complex, capturing both stable patterns and transient, spontaneous events (Cabral *et al*., 2022). Interpreting EEG is challenging because it involves distinguishing meaningful neural activity from noise and artifacts while accounting for the brain’s intricate spatial and temporal dynamics. Given these challenges, clinical interpretation of EEG can greatly benefit from machine-driven augmentation, enabling the identification of novel or hidden features through AI methodologies.

The emergence of big data approaches in neuroscience has underscored the importance of routine EEG as a high-dimensional, multi-modal dataset that can be analyzed in conjunction with other types of electronic health records (Josephson *et al*., 2024). When integrated with demographic parameters (e.g., age, sex), expert neurological reports, diagnostic classifications, and therapeutic interventions, EEGs facilitate a comprehensive, data-driven exploration of brain health across populations (Cassani *et al*., 2018). Unlike controlled laboratory EEG studies, routine clinical EEGs reflect real-world patient variability, making them highly valuable for biomarker discovery, disease progression modeling, and treatment response prediction (Lemoine *et al*., 2024).

### Clinical challenges in EEG evaluation

Interpreting EEG presents multiple challenges that impact diagnostic accuracy, clinical decision-making, and research applications (Greenblatt, Beniczky and Nascimento, 2023). In general, brain signals have a high inter-individual variability (Marek *et al*., 2022). Variations in brain anatomy, age, and medication use further contribute to this complexity. Also, some benign epileptiform variants appear in healthy individuals, making it difficult to distinguish normal activity from abnormalities (Benbadis and Lin, 2008).

EEG analysis relies heavily on visual inspection by clinicians, introducing inter-rater variability (Beniczky *et al*., 2017; Jing *et al*., 2020). Differences in training and experience influence the consistency of EEG reports, particularly in identifying epileptiform discharges and non-specific abnormalities (Tatum, 2012). Borderline or ambiguous patterns, such as slow waves or small spikes, may receive inconsistent classifications across evaluators (Jing *et al*., 2020). Furthermore, routine EEG recordings contain artifacts from eye movements, muscle activity, cardiac signals (ECG), and external electrical sources (Tatum, 2013a). Factors such as patient movement, poor electrode contact, and sweating can introduce additional distortions.

The limited duration of routine EEG, typically 20–60 minutes, poses further diagnostic challenges (Reardon *et al*., 1999). Infrequent epileptiform discharges or paroxysmal events may not be captured, reducing the sensitivity of EEG in epilepsy diagnosis. The risk of false positives also increases when normal variants are misclassified as pathological patterns (Benbadis and Kaplan, 2019). Moreover, differences in EEG systems, electrode placements, and software configurations across hospitals hinder cross-site comparisons. Variations in montages, filter settings, and gain adjustments affect signal visualization, further complicating standardization.

These challenges contribute to variability in EEG interpretation and increase the likelihood of misdiagnoses, particularly in patients with non-specific symptoms (Tatum, 2013b). Addressing these issues through standardized protocols and automated analysis methods is crucial for improving the reliability of clinical EEG evaluation (Carlson, 2015).

### Objectives

We aim to prepare a large dataset of routine clinical EEG recordings for research, featuring multiple versions of pre-processed EEG data. These recordings will be standardized in a common brain space, defined by anatomical and functional landmarks, and accompanied by clinically relevant labels. Our ultimate goal is to enhance the electrophysiological evaluation of patients by facilitating peer review and improving risk stratification for newly acquired EEG recordings. We also seek to minimize biases by integrating expertise from multiple specialists and prototyping a decision-support system that leverages collective intelligence for EEG interpretation.

### Experimental design

This protocol details a multi-center, retrospective, and observational method for analyzing population-level EEG data. By incorporating data from multiple hospitals, the protocol ensures the inclusion of a diverse patient cohort, which increases the generalizability of findings to real-world clinical settings. This multi-center approach necessarily introduces heterogeneity in data acquisition, including variations in EEG hardware, recording protocols (e.g., reference electrode placement), and technician expertise. We account for this variability during data processing. Furthermore, this inherent heterogeneity strengthens the protocol’s translational relevance, as it reflects the conditions under which a new technique would be clinically implemented.

The protocol outlines the processing of existing EEG recordings that were acquired during routine diagnostic evaluations. These recordings were originally performed for clinical indications, such as diagnosis of paroxysmal events or the assessment of encephalopathy and altered mental status. The use of routine clinical data, which often includes standard activation procedures like hyperventilation and photic stimulation, ensures that the protocol is applicable to standard medical practice. As the design is observational, it introduces no new treatments or interventions. Instead, it focuses on the association between patients’ EEG patterns and their documented clinical features and outcomes. We retrospectively define clinical subgroups based on diagnostic criteria and neurological assessments from the original EEG reports.

A key strength of this protocol is its use of population-level data. The method encompasses all inpatient and outpatient EEG examinations conducted within specific public hospitals over a defined period. This comprehensive inclusion criterion minimizes selection bias. Consequently, analyses based on this protocol should accurately reflect the full spectrum of EEG studies and patient populations encountered in routine clinical practice.

### Development of the protocol

We have developed this protocol to standardize the analysis of routine clinical EEG recordings collected from multiple public hospitals within the health authority of British Columbia. Although hardware and software are harmonized within the authority at any given time, these systems continue to evolve. Consequently, the primary challenge is managing data heterogeneity that arises from longitudinal changes in equipment, variations in electrode configurations (e.g., reference and ground placement), differences in technician expertise, and other environmental factors. To address this variability, we developed a standardized pre-processing pipeline. This pipeline renders the data suitable for large-scale research, facilitates comparative studies within a single health system, and promotes interoperability for international collaborations.

While the individual steps within our protocol are standard in EEG analysis, their integration into a robust, automated pipeline is critical for processing large-scale clinical data. Unlike data from controlled laboratory experiments, our retrospective dataset precludes manual inspection and necessitates a minimalistic yet robust workflow capable of handling significant heterogeneity. These challenges include variability in channel nomenclature, inconsistent recording durations, and the sporadic presence of clinical activation procedures, such as hyperventilation and photic stimulation, or periods of isoelectric signal. Furthermore, analyses must be adapted to the constraints of a limited-channel clinical montage. Our protocol is therefore designed not for the novelty of its individual components, but for its essential function: to systematically harmonize these disparate recordings on a large scale. This automated standardization is a prerequisite for ensuring that downstream analyses accurately reflect genuine neurophysiological variance rather than technical artefacts, thereby enabling the discovery of valid biomarkers from real-world clinical data.

The protocol begins with the conversion of raw EEG data from a proprietary manufacturer format into a standardized structure (EDF+). We then apply a series of pre-processing steps, including artifact rejection, re-referencing to a common average, and spatial normalization. These procedures reduce the impact of instrumental and environmental variance, thereby improving the reproducibility and generalizability of subsequent analyses. While the current protocol focuses on EEG pre-processing, it is designed to be extensible. Future work will integrate associated clinical information, including EEG reports, diagnoses, and medication profiles. Integrating these data with the pre-processed EEG recordings provides a powerful resource for biomarker discovery and the development of predictive clinical models.

We validated the utility of this protocol by applying it to several studies of EEG-derived “brain age”, a biomarker of brain health and cognitive status. One study compared EEG signal decomposition techniques for predicting chronological age and found that stationary component analysis yielded higher accuracy than conventional frequency-band filtering (Paliwal *et al*., 2024). A second study treated EEG signals as symbolic sequences and identified recurring motifs of neural activity that predicted age with an accuracy comparable to that of traditional spectral methods (Klymenko *et al*., 2023). Finally, our research on the brain age gap, which is the difference between chronological and biological age, challenged the hypothesis that its variance increases with age due to stochastically accumulated pathology. Instead, our findings suggest that factors present earlier in life may be more determinant of an individual’s brain age trajectory (Kalapun *et al*., 2024). These validation studies demonstrate that the standardized dataset can support sophisticated neurophysiological analyses and generate novel clinical insights.

### Applications of the method

The availability of large-scale, population-based routine clinical EEG data presents transformative opportunities for clinical neuroscience, population neuroscience, and artificial intelligence (AI)-driven brain research. These datasets integrate demographic, diagnostic, treatment-related and expert knowledge, enabling various scientific investigations.

Routine clinical EEG data can facilitate the studies of age-related changes in neural activity across the lifespan. By analyzing developmental and ageing-related alterations in neural oscillations, we can establish natural links between early and late life stages (Di Biase *et al*., 2023). These data would also allow us to characterize age-related trajectories in brain parameters and identify trajectories associated with specific clinical conditions. Considering diverse clinical groups, defined by established diagnoses and expert knowledge (EEG reports), further enhances the scope of these investigations.

Our dataset also supports large-scale research in population neuroscience. Unlike experimental sample studies, which often involve highly controlled conditions, routine clinical EEG is collected in real-world clinical settings, minimizing selection bias. Such an approach broadens the generalizability of findings and enables unbiased investigations of brain function across diverse populations and clinical subgroups (Li *et al*., 2024). Furthermore, where medication records are available, this dataset can help explore inter-individual variability in brain function and its modulation by pharmacological interventions.

Standardization and source localization are essential to enhance the interpretability of scalp-recorded EEG signals. Mapping EEG data onto a common anatomical space (e.g., the Destrieux atlas) or functional networks (e.g., resting-state fMRI parcellations) reduces ambiguity in interpretation (Destrieux *et al*., 2010; Thomas Yeo *et al*., 2011; Schaefer *et al*., 2018). Reconstructing neural source dynamics and quantifying the variability across regions of interest within an atlas could facilitate multi-modal neuroimaging research. This approach allows for the integration of EEG with MRI and cellular studies, enhancing research capabilities even in the absence of complete multi-modal datasets from the same individuals (Lotter *et al*., 2024).

In the field of AI, large-scale EEG datasets provide a foundation for developing advanced machine-learning models. The availability of extensive data enables the creation of foundation models for EEG, which employ vast amounts of data to generate general-purpose representations (Liang *et al*., 2024). In turn, extracting learned features from EEG time series instead of relying solely on manually engineered EEG features may improve classification accuracies. Given the naturally occurring labels in clinical EEG data, such as age, sex, and spatial information, these models can be trained using self-supervised and supervised learning approaches to maximize their adaptability and efficiency.

Finally, EEG synchronized with external time-dependent variables (e.g., geomagnetic activity or atmospheric pressure fluctuations) offers new avenues for investigating brain-body interactions. Integrating EEG with environmental and physiological data could provide further insights into the influence of external factors on brain activity and cognitive function (Sulentic *et al*., 2023).

## Materials

### Patients’ electronic health records

We identify approximately 42,000 EEG studies conducted for diagnostic purposes at four public hospitals within a health authority in British Columbia. These EEG recordings span approximately eight consecutive years of clinical practice. Selection criteria are minimal and primarily require the ability to link EEG data with patients’ medical records and corresponding EEG reports. Neurologists affiliated with the participating hospitals evaluated all EEG recordings during diagnostic workup. The dataset includes both inpatients and outpatients, with an approximate ratio of 3:1 favouring outpatients. Patient ages range from one year to over 100 years.

Additional clinical data are available for EEG from inpatients, including disease classification based on ICD-10 codes and diagnostic categorization by type (most responsible, secondary, pre-admission, and post-admission comorbidities). The inpatient cohort is further categorized using a patient classification system developed by the Canadian Institute for Health Information (CIHI). Specifically, we rely on the Case Mix Group (CMG) methodology, widely implemented in Canadian public hospitals as part of electronic health records (*Case Mix Group+ (CMG+) Directory*, 2022). CMG assigns hospitalized patients to clinically meaningful groups based on primary diagnosis, procedures performed, comorbidities, complications, age, sex, and discharge status. These groups are further organized into 21 major clinical categories, which provide a broader classification framework.

Mental disorders represent one of these major clinical categories and include several sub-categories. CMG groups within the mental disorders category encompass conditions such as schizophrenia, schizotypal disorder, bipolar disorder, depression, and dementia. Compared to ICD-10 diagnostic codes, CIHI’s CMG system offers a higher-level classification that facilitates the subtyping of inpatients based on clinical and resource utilization characteristics.

### Routine Clinical EEG recordings

We obtain EEG recordings from four hospitals, where all EEG units use harmonized Natus hardware and firmware. Each acquisition station is equipped with a Natus EEG32U amplifier that records from 20 scalp electrodes positioned according to the international 10–20 system. The standard setup also includes two dedicated electrooculography (EOG) channels (Pg1, Pg2), two linked-ear reference channels (A1, A2), and eight auxiliary (AUX) channels. Technicians typically use the AUX channels to record electrocardiography (ECG) and, in some cases, additional physiological signals. While the hardware is standardized, a significant source of variability arises from the technician-dependent choice of the recording reference. The acquisition software permits the selection of several reference schemes, including the linked-ear electrodes (A1/A2), a designated auxiliary channel, or a system-defined reference. Because this selection is not standardized across recordings, the reference configuration is expected to be consistent within any single file but vary across the dataset.

Each routine clinical EEG primarily captures spontaneous, resting-state brain activity. The protocol also includes two standard activation procedures to enhance diagnostic yield: hyperventilation (HV) and intermittent photic stimulation (IPS). The HV procedure involves 2–4 minutes of deep, rapid breathing, while the IPS procedure consists of presenting a stroboscopic light flashing at frequencies ranging from 1 to 60 Hz. The beginning and end of both HV and IPS periods are annotated in the digital recording files, which allows for their precise identification during subsequent analysis.

### Procedure

#### Overview

Our protocol consists of four main stages: (1) standardization, (2) segmentation, (3) correction for artifacts, and (4) brain source reconstruction. This combination ensures data consistency, quality, and interpretability.

In the standardization stage, we de-identify the data, filter and resample it to a common sampling rate and frequency band, and align it to an *a priori* defined set of EEG channels. This step ensures the first approximation to uniformity across recordings, regardless of the original acquisition settings.

During segmentation, we extract up to three types of EEG segments based on availability: a relatively long segment of spontaneous activity and intervals corresponding to intermittent photic stimulation (IPS) and hyperventilation (HV). These segments capture different physiological states, facilitating a more comprehensive analysis of EEG dynamics.

In the pre-processing stage, we focus on improving data quality by identifying and removing bad channels and eliminating common artifacts. This step enhances signal integrity and reduces noise, ensuring that subsequent analyses rely on cleaner data.

Finally, after brain source reconstruction, we interpret EEG signals in terms of neural activity within specific regions of interest (ROIs). These ROIs are defined using widely recognized cortical parcellations. This process transforms the original EEG recordings into a normalized, dimensionless form referenced to standard anatomical and functional landmarks. As a result, the reconstructed data becomes independent of EEG hardware, montage, channel configuration, and other acquisition-specific factors, enabling more reliable cross-study comparisons.

#### Initial conversion and de-identification of EEG records

EEG system manufacturers use proprietary data formats, which can complicate data sharing and analysis. In this study, EEG data were originally recorded in Natus’ proprietary format. To facilitate interoperability, we convert the raw data into the European Data Format plus (EDF+), a widely used standard for storing and exchanging biomedical time series. This conversion is performed using EDFExport.exe, utility distributed with Natus Neuroworks software.

The EDF format assumes continuous recordings. However, EEG technicians may start and stop EEG recordings multiple times during a session, such as when testing and adjusting electrode impedances. As a result, during conversion to EDF, discontinuous EEG segments are linked using digital zeros, which appear as artificially small fluctuations in EEG amplitude. These inserted zeros indicate gaps in the recording and should be considered when analyzing the data.

Following conversion to EDF, we apply a de-identification process to protect patient privacy. We differentiate between de-identification and anonymization. De-identification removes personally identifiable information while still allowing authorized researchers to link EEG data to electronic health records (EHRs) when necessary. In contrast, anonymization would eliminate any possibility of re-identification. To perform de-identification, we remove fields containing sensitive information about the patient, EEG technician, or neurologist. This process is implemented using PyEDFlib, a Python package providing an interface for EDFlib, a C/C++ library designed to read and write EDF+ files.

#### Selection of EEG channels

Before processing each EEG record, we first verify the presence of a mandatory set of channels, including Fp1, Fpz, Fp2, F3, F4, F7, F8, Fz, T3, T4, C3, C4, Cz, T5, T6, P3, P4, Pz, O1, and O2. This set aligns with the International 10-20 system, with some modifications. We included Fpz, as it is part of the Natus EEG32U amplifier. This mandatory set is chosen to maximize the number of EEG records retained for analysis. We exclude EEG records that lacked one or more of these required channels.

In addition to the mandatory channels, we include optional channels such as A1 and A2, and recordings for EOG and ECG. However, we encounter inconsistent channel naming conventions across medical facilities. These variations extend beyond simple case differences (e.g., “fp1” vs “FP1”). For example, the left EOG channel appears under multiple names, including “EOG1,” “EOG 1,” and “L EOG”. Similarly, the right EOG channel can be labeled as “EOG2,” “EOG 2,” or “R EOG”. Empirically, we determine that EOG channels are commonly stored under the following names: “EOG 1,” “EOG 2,” “EOG1,” “EOG2,” “L EOG,” “R EOG,” “PG1,” and “PG2”. Likewise, ECG channels appear under names such as “ECG1,” “ECG2,” “EKG,” “EKG1,” and “EKG2”.

Still certain channel names remain unrecognized and need to be changed. For example, we rename “L EOG” to “EOG1,” “R EOG” to “EOG2,” and “EKG” to “EKG1” to standardize the channel labels. Also, the EDF files produced after conversion from the proprietary Natus format lack explicit channel-type information. We designate four types of channels: EEG, EOG, ECG, and MISC (miscellaneous). More specifically, we explicitly assign all mandatory channels to the “EEG” category. We classify EOG and ECG channels accordingly. We do not include the channels A1 and A2 as EEG type data channels. We label these channels as “MISC”. All other channels are discarded.

Following channel standardization, we apply a uniform filtering and resampling protocol. We first exclude long-term recordings with low native sampling frequencies (e.g., 200 Hz). For all other recordings, we begin by applying a 60 Hz notch filter (with a width of 0.3 Hz) to all EEG, EOG, and ECG channels to remove power line interference. Next, we band-pass filter the EEG channels between 0.5 Hz and 55 Hz. For both filtering steps, we use a fourth-order Butterworth filter applied in both the forward and backward directions to ensure zero-phase distortion. Finally, we resample all recordings to a uniform sampling frequency of 256 Hz.

#### Segment Extraction: HV, IPS, and Resting-State EEG

We extract three types of segment from each EEG record: hyperventilation (HV), intermittent photic stimulation (IPS), and resting-state EEG, upon availability. Using the annotations stored in the EDF files, we identify HV and IPS intervals. To account for transient effects, we add a 30-second buffer before and after each segment. The extracted HV and IPS segments, if present, are saved as separate EDF files, preserving the original annotations to enable precise event timing identification when needed.

For resting-state EEG, we select a 360-second segment that meets the following criteria. We exclude intervals containing flat signal segments (digital zeros), optical stimuli (IPS), or hyperventilation activation (HV). Also, we disregard the first 420 seconds of each recording to avoid instrument setup artifacts. More specifically, we define flat intervals as segments where the absolute voltage difference between consecutive samples remained below 1 µV for at least 10 seconds. Any identified flat, IPS, or HV segments receive an additional 30-second buffer before and after their boundaries. We then save the first 360-second segment that satisfied the above conditions as a separate EDF file. All extracted HV, IPS, and resting-state EEG segments are stored individually, with corresponding channel type and filtering parameter information included in the metadata.

#### Further pre-processing and correction for artifacts

The previous filtering and segmentation steps produce IPS, HV and resting-state EEG recordings with standardized length, sampling frequency, data bandwidth, and channel set. These steps also remove gross artifacts, such as flat intervals and power line interference. However, no additional quality enhancements are applied at this stage. Specifically, we did not identify noisy channels, remove ocular and cardiac artifacts, or address variability in EEG referencing across different medical facilities. The primary objective of the next pre-processing step was to refine the dataset by addressing these issues and improving data quality for subsequent analyses.

To standardize EEG pre-processing, we apply PREP (Pre-processing Pipeline for EEG), an automated pipeline designed to detect and correct common EEG artifacts in a robust and reproducible manner (Bigdely-Shamlo *et al*., 2015). PREP is widely used to pre-process EEG data prior to further analysis. We apply PyPREP, a Python implementation of this pipeline. Pre-processing involves four sequential stages: drift removal, line noise removal, detection of noisy (bad) channels, and calculation of a robust average reference using clean electrodes. Although we had already removed power line noise and drift artifacts in the previous pre-processing step, we retained these procedures as part of the standard PREP workflow for consistency.

To further standardize the EEG recordings, we re-reference the signals to the grand average reference. A key challenge in this process is the exclusion of noisy channels from the reference calculation. PREP applies an iterative approach, updating both the grand average reference and the set of bad channels until convergence. Once finalized, bad channels are either interpolated (when sufficient clean data is available) or marked as unusable if interpolation is not feasible. This procedure is expected to improve the signal-to-noise ratio (SNR) of the EEG data.

Using PREP’s default criteria, identifying bad channels is common. As a result, despite enforcing a mandatory set of channels, some EEG recordings lack one or more of these required channels. This limitation contributes to our rationale for transforming EEG data into the source space, as discussed in a later section.

Following the PREP pipeline, we apply Independent Component Analysis (ICA) to remove ocular and cardiac artifacts (Hyvarinen, 1999). Specifically, we use the Fast ICA algorithm as implemented in MNE-Python (Gramfort *et al*., 2014). Before ICA decomposition, EOG and ECG channels are filtered using zero-phase finite impulse response (FIR) filters within specific frequency bands: 1–5 Hz for EOG and 8–16 Hz for ECG. These filtered signals are retained in the output to facilitate artifact identification. No additional filtering is applied to EEG sensor channels before ICA processing. To remove physiological artifacts, the MNE algorithm identifies ICA components that exhibit strong correlations with the filtered EOG and ECG signals and exclude them from the EEG data. We use default ICA parameters as specified by MNE-Python.

We store the processed EEG records in .fif format, the native format of MNE-Python. This format is chosen because it preserves metadata related to pre-processing steps and seamlessly integrates with subsequent processing stages, which are also conducted using MNE-Python.

#### Reconstruction of EEG source dynamics

Our analysis assumes that all neural sources are confined to the cortical surface, whose shape is defined by a template. We also adopt the standard assumption that all brain sources can be represented as current dipoles with arbitrary orientations that remain constant over time. Within the MNE Python software framework, this assumption translates to using a surface-based source space with unconstrained dipole orientations.

Source reconstruction requires both the patients’ anatomical MRI and EEG sensor positions on the head. However, clinical EEG recordings rarely include this information. To address this limitation, we use a template MRI “fsaverage” derived from the average of 40 brains after normalization to a standard space and distributed with FreeSurfer, along with standardized 10-20 sensor locations (Fischl, 2012).

We reconstruct source dynamics for predefined regions of interest (ROIs). One approach uses ROIs defined according to the Destrieux atlas, an anatomical cortical parcellation commonly used in MRI-based neuroimaging (Destrieux *et al*., 2010). This atlas segments the cortical surface into 74 regions per hemisphere (148 total) based on sulcal and gyral anatomy, further grouped into six major brain lobes.

In addition to the anatomical parcellation, we incorporate a functional cortical parcellation using the Schaefer atlas. This atlas, developed for resting-state functional MRI (rsfMRI), segments the cerebral cortex into network-based regions aligned with large-scale intrinsic connectivity networks (Thomas Yeo *et al*., 2011). We use the 50-ROI per hemisphere parcellation, which assigns each ROI to one of seven functional networks derived from rsfMRI data (Schaefer *et al*., 2018).

Each region of interest (ROI) covers a certain area of the cortical surface and contains multiple dipolar sources defined by FreeSurfer’s “fsaverage” cortical surface. We compute forward solutions (lead fields) for each cortical source using the MNE-Python framework (Baillet, Mosher and Leahy, 2001; Gramfort *et al*., 2014). This computation relies on a three-layer boundary element model (BEM) of the head, with standard conductivity values of 0.3, 0.006, and 0.3 S/m assigned to the inner skull, outer skull, and scalp layers, respectively.

To reconstruct temporal source activity, we apply a scalar single-source minimum variance beamformer (Frost, 1972; Gramfort *et al*., 2014; Ilmoniemi and Sarvas, 2019). This method estimates source activity while suppressing interference from other brain regions and external noise. To mitigate depth-related biases, we normalize the reconstructed time courses using the root mean square (RMS) amplitude of sensor level noise projected to the source locations using beamformer weights. This normalization produces dimensionless pseudo-Z units (Van Drongelen *et al*., 1996; Westner *et al*., 2022), where pseudo-Z represents the ratio of the reconstructed signal to the projected noise. However, the pseudo-Z amplitudes of source waveforms may still vary considerably across EEG recordings due to differences in overall noise levels.

To prevent statistical biases, we apply an additional normalization to all reconstructed time courses. This is done using the square root of a global power ratio, defined as the ratio of signal power summed over all electrodes to the total noise power for the given recording. This second normalization ensures that source time courses from different recordings have comparable orders of magnitude. The corresponding normalization constant is stored with each recording, allowing the original pseudo-Z magnitudes to be reconstructed if necessary.

We extract a single representative time course for each ROI by applying Principal Component Analysis (PCA) to the time courses of all dipolar sources within the ROI. The first principal component, which explains the most of variance across these sources, is selected as the final ROI time course.

We store the ROI-specific neural dynamics in the Hierarchical Data Format (.hdf5), a flexible scientific file format commonly used in machine learning and high-performance computing. Each HDF5 file contains the ROI-specific EEG time series, along with relevant metadata, including ROI names, spatial coordinates of each ROI’s center of mass, beamformer weights associated with each ROI, and a global sensor-level pseudo-Z value computed for the entire duration of EEG recording. Source reconstruction is performed twice, separately for the anatomical and functional parcellations, for all available EEG segments, and separately for HV, IPS, and spontaneous activity.

#### Overview of Software applied

All processing and analysis are performed in Python, leveraging a suite of open-source neuroinformatics packages. We first convert the raw data from the proprietary Natus format into the European Data Format Plus (EDF+), a widely adopted standard for clinical electrophysiology <https://www.edfplus.info/>. We use the PyEDFLib library for this conversion and to programmatically de-identify patient information contained within the file headers <https://github.com/holgern/pyedflib>. Initial robust pre-processing, including the automated detection and interpolation of bad channels, is performed using the PyPREP pipeline <https://github.com/sappelhof/pyprep>.

Subsequent pre-processing and analysis stages are conducted within the MNE-Python environment <https://mne.tools/stable/index.html>. MNE-Python provides a comprehensive toolkit for EEG/MEG analysis, from filtering and artifact correction to statistical inference and visualization. Throughout these stages, we store the data in MNE-Python’s native FIF format. This format retains the processed data along with all associated metadata, such as channel locations, event markers, and the complete processing history, which ensures full reproducibility of our workflow.

For source reconstruction, we generate anatomical head models using Freesurfer and its templates <https://freesurfer.net>. We then apply a beamforming algorithm, which is implemented as an in-house Python library, to estimate source-level activity. The resulting source-space time series are large numerical arrays, which we stored in the Hierarchical Data Format (.hdf5) <https://github.com/HDFGroup/hdf5>.

### Anticipated results

#### Overview

Our neuroinformatic framework organizes all EEGs into up to three neurophysiological states: spontaneous activity, hyperventilation, and photic stimulation, depending on availability. Each state is represented at multiple levels of data processing, providing a structured methodological hierarchy.

At the first level, EEG signals remain in their original sensor space, closely matching the recordings evaluated by neurologists. The second level involves re-referenced EEG data with bad channels removed, and corrected for the eye (EOG) and cardiac (ECG) artifacts, improving signal quality. The third level consists of source-reconstructed activity, mapped onto functional and anatomical cortical parcellations. This source-level representation serves as the foundation for further analyses.

Due to the limited number of EEG electrodes, the effective spatial resolution of source reconstruction remains constrained. Consequently, our analyses will primarily focus on the temporal properties of neural dynamics rather than functional connectivity, which typically examines coordinated activity across distinct brain regions. The transformation from sensor space to source space essentially represents the upsampling of spatial resolution by projecting EEG signals onto a standardized brain space.

Beyond EEG time series, data harmonization extends to clinical metadata. Each EEG, which is linked to its corresponding neurological report, and for inpatients, their classification is based on established diagnoses. We process EEG reports to extract structured clinical evaluations using a predefined schema, ensuring consistency and facilitating large-scale analyses.

#### Illustrative example

To illustrate the pre-processing steps described in previous sections, we present EEG data at both the sensor and source levels from a single participant with a typical resting-state EEG recording. Figure 1 displays EEG signals from a 20-second interval extracted from the original data segment. A total of 25 channels are available, including 19 EEG sensor channels, resulting in an initial EEG data rank of 19.

**Figure 1.**
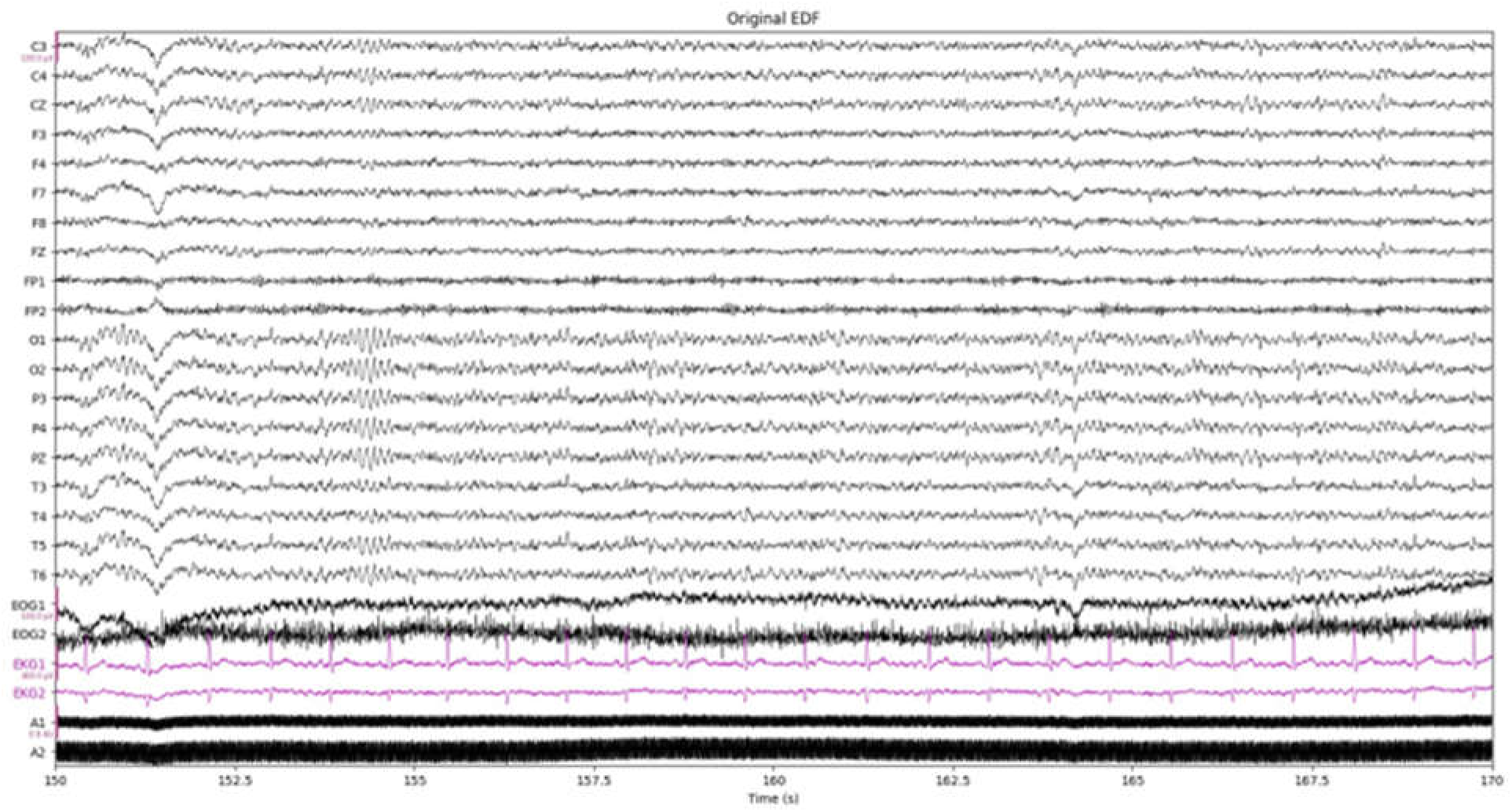
*Illustrative short EEG segment at the sensor level*. The figure shows an original 20-second EEG segment from a single participant’s resting-state recordings, relative to the study reference.

The subsequent PREP pre-processing pipeline involves re-referencing the EEG sensors, discarding one channel due to high-frequency interference, and interpolating two additional channels. The ICA algorithm removes two components that exhibit high correlations with EOG signals. However, in the given EEG, ICA fails to confidently identify and remove components linked to ECG artifacts. After completing all pre-processing steps, but before source reconstruction, the effective data rank is reduced to 15.

Figure 2 presents the power spectra of EEG signals before and after pre-processing, which is estimated based on the entire six-minute segments. Both spectra exhibit a typical 1/f distribution, with a prominent peak in the alpha frequency range. The pre-processed signals appear visually cleaner, with substantial attenuation of noise artifacts. In particular, interference at around 20 Hz is nearly eliminated, and artifacts at around 40 Hz are significantly reduced. Also, the pre-processed data show evidence of a beta-band peak, observed as a deviation from the expected 1/f trend.

**Figure 2.**
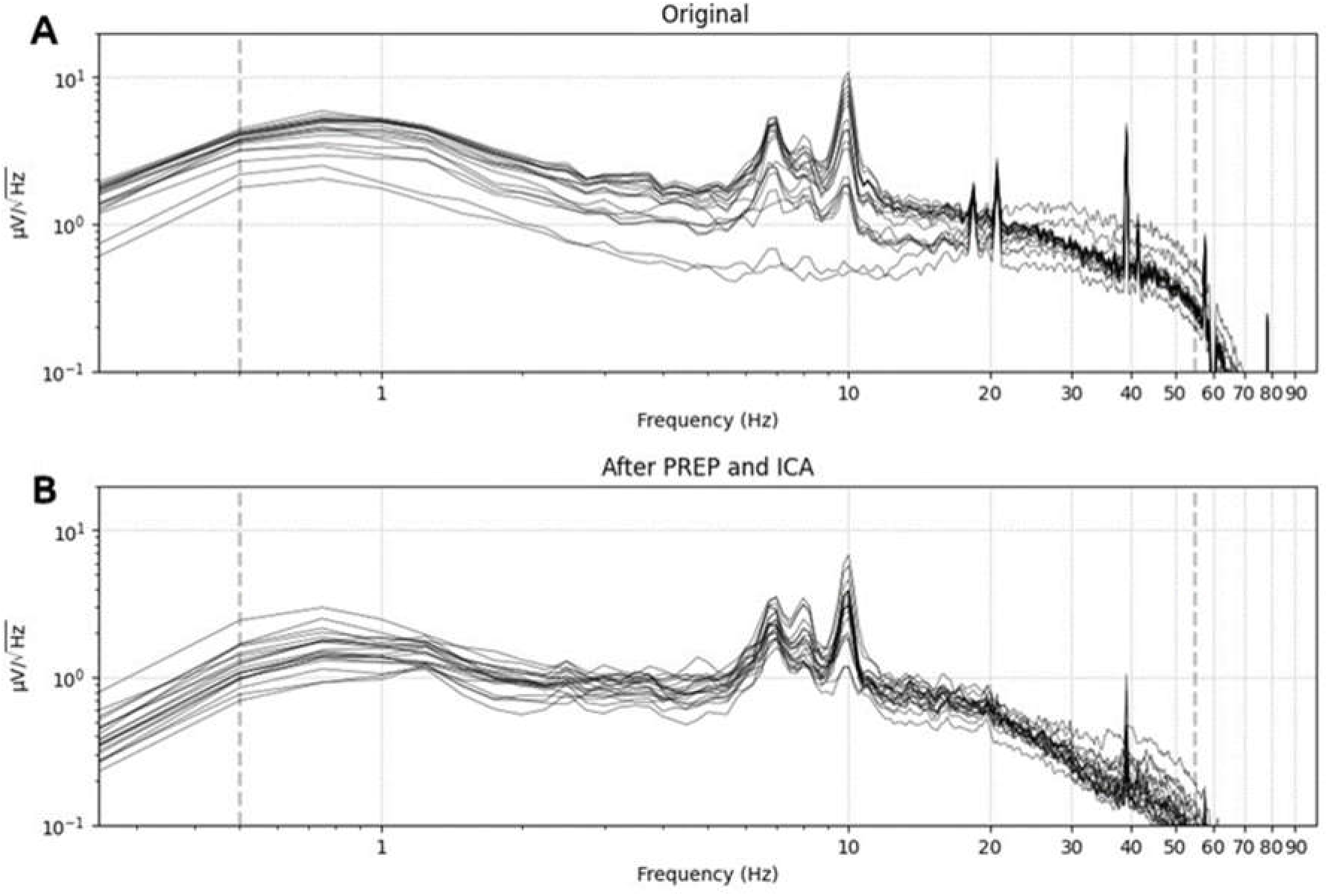
*Power spectra of EEG signals before (A) and after (B) pre-processing*. Individual curves represent individual channels. Both spectra follow a typical 1/f distribution with a prominent alpha peak. Pre-processing reduces noise artifacts, nearly eliminating interference around 20 Hz and attenuating artifacts near 40 Hz. The cleaned data also reveal a beta-band peak, deviating from the expected 1/f trend.

After source reconstruction, EEG signals are mapped to a set of regions of interest (ROIs), each represented as a time series in dimensionless pseudo-Z units. The Destrieux anatomical parcellation includes 148 ROIs, while the Schafer functional parcellation contains 100 ROIs. To illustrate the spatial distribution of EEG activity, we computed the mean spectral density of alpha-band activity (8–12 Hz) for each ROI. Figure 3 visualizes this metric by projecting the estimated values onto an inflated brain surface. The strongest alpha-band power is concentrated in the occipital lobe, consistent with expected physiological patterns. However, the spatial accuracy of these results is constrained by the limited rank of the data. Also, we assign a single alpha power value to each ROI, though the actual distribution within an ROI may be non-uniform.

**Figure 3:**
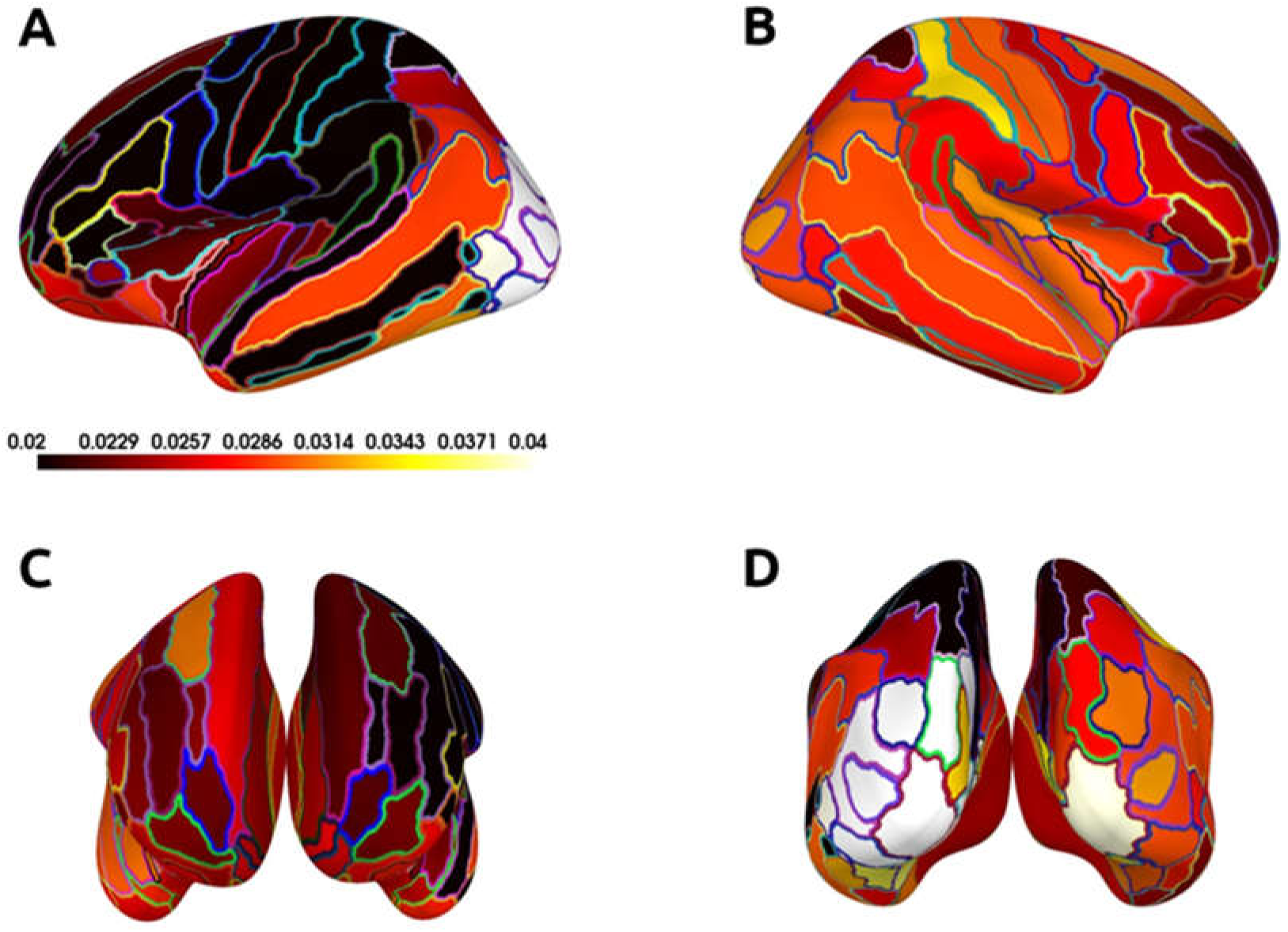
Spatial distribution of resting-state alpha-band spectral power. The figure displays the source-localized power spectral density in the alpha band (8–12 Hz) across the 148 regions of the Destrieux atlas, estimated from a representative resting-state EEG recording. The power is mapped onto an inflated cortical surface, showing the left (A) and right (B) hemispheres, along with rostral (C) and caudal (D) views. Solid lines delineate the boundaries of each region of interest. Consistent with established neurophysiology, the highest concentration of alpha power is localized to the occipital lobe. The color bar indicates power spectral density in arbitrary units (a.u.).

### Limitations and opportunities

#### General considerations

Our pre-processing strategy is guided by a fundamental trade-of. Each procedure, while intended to improve data quality by removing artifacts such as those from eye movements, also risks introducing uncontrolled distortions or removing genuine neural signals. Given this constraint, and because our large-scale dataset precludes manual intervention, we adopted a relatively minimalistic approach designed to balance artifact suppression with the preservation of underlying data integrity. This conservative strategy is particularly important for subsequent source reconstruction, which may be sensitive to distortions in the data’s spatial and temporal structure.

The selection of this approach is also informed by the absence of a single, universally accepted metric to define optimal data quality. This represents a challenge that underscores the need for principled, rather than overly aggressive, automated processing pipelines (Delorme, 2023).

A potential limitation of our protocol is the absence of explicit annotations distinguishing between eyes-open and eyes-closed periods within the resting-state recordings. This distinction is neurophysiologically critical, as the state of the eyes is the primary modulator of the posterior dominant alpha rhythm. We cannot rule out brief and spontaneous periods of eye opening or eye closing. However, this concern is partially mitigated by the standard clinical procedures used during data acquisition. At the beginning of each session, technicians are expected to routinely assess EEG reactivity by instructing the patient to open and close their eyes. Consequently, it is highly probable that the subsequent segments of spontaneous activity, which constitute the data for our analysis, were recorded while the patient was in a relaxed, eyes-closed state.

#### Reconstruction of EEG source dynamics

Reconstructing brain source dynamics from sensor-level recordings becomes a standard approach in high-density EEG (Westner *et al*., 2022). This process involves solving the bioelectromagnetic inverse problem, which aims to estimate brain activity from electrical or magnetic signals recorded outside the skull. Due to the ill-posed nature of this problem, no unique solution exists without making specific assumptions about the properties of brain sources. While extensive research addresses this topic, a detailed discussion is beyond the scope of this paper.

Pre-processing steps such as re-referencing, bad channel interpolation, and independent component analysis (ICA) artifact removal reduce the data rank, or effective number of independent channels in the EEG study. Despite the relatively low number of EEG channels, we argue that source reconstruction remains beneficial, providing several advantages without introducing evident signal degradation.

First, transforming EEG data from sensor space to a standardized brain space mitigates the problem of missing data arising from variations in electrode configurations. Even when restricting analyses to a mandatory set of channels, some electrodes may be unavailable due to pre-processing steps.

Second, using a standardized anatomical or functional space facilitates the integration of EEG data collected with different electrode placements and channel configurations. This is particularly relevant when incorporating clinical EEG datasets from multiple medical centres, where recording protocols may follow different conventions, such as the modified Maudsley system or cap-based recordings (Seeck *et al*., 2017).

Third, mapping EEG data to an anatomical space (e.g., the Destrieux cortical atlas) or a functional space (e.g., the Schaefer parcellation) enhances the interpretability of further results (Destrieux *et al*., 2010; Thomas Yeo *et al*., 2011; Schaefer *et al*., 2018). This approach facilitates the comparison of specific effects across brain regions.

Fourth, aligning EEG data with other brain parameters, including neuroimaging findings and cellular-level research, enables multi-modal integration. Even when multi-modal data are unavailable within the same cohort, using a common reference space supports cross-study comparisons and broader neuroscientific insights (Lotter *et al*., 2024).

Finally, source reconstruction techniques, such as beamforming, provide additional interference suppression by attenuating non-dipolar spatial patterns (Nazarpour *et al*., 2007; Metsomaa *et al*., 2024). This further enhances signal quality and improves the accuracy of EEG-based functional mapping.

#### Comparison with other methods

Our study protocol builds upon the foundations laid by existing large-scale EEG database initiatives, yet it introduces a distinct methodological approach. We design our data processing and analysis framework by comparing it with three established research resources: the Temple University Hospital (TUH) EEG Corpus (Obeid and Picone, 2016), the Harvard Electroencephalography Database (HEEDB) (Sun *et al*., 2025), and the SCORE database (Beniczky *et al*., 2017; Tveit *et al*., 2023). Each of these corpora provides a valuable but different model for data collection, annotation, and accessibility.

The first major initiative, the TUH EEG Corpus, represents an extensive, publicly available collection of routine clinical EEG data from Temple University Hospital (Harati *et al*., 2014; Obeid and Picone, 2016). This ongoing project has curated and organized nearly a decade of recordings. A key feature of the TUH-EEG corpus is that a subset of records is paired with the original clinician reports. The data are structured into multiple sub-datasets, each emphasizing specific clinical tags. These tags range from macro-labels that classify entire recordings, such as “normal” or “abnormal,” to precise annotations of specific electrographic events, including artifacts and epileptiform discharges.

Two other prominent databases, HEEDB and SCORE, present alternative approaches. HEEDB is notable for its immense sample size and the availability of raw EEG data for approved research projects (Sun *et al*., 2025). However, their database metadata primarily includes only patient age and gender https://bdsp.io/content/harvard-eeg-db/4.1/, although the clinical context for these recordings is extensive. In contrast, the SCORE database consists of EEG recordings collected from clinical users of the hiSCORE software (Holberg EEG, Bergen, Norway) and is available only to participating researchers (Beniczky *et al*., 2017; Tveit *et al*., 2023). Expert physicians label these recordings, which are integrated with extensive metadata, including links to full EEG reports and specific clinical findings. Similar to the SCORE initiative, and unlike the TUH-EEG corpus, the data in our project are not publicly available.

Our methodology diverges from these precursors in several key aspects, primarily through our emphasis on comprehensive data harmonization. The central goal of our project is to develop large-scale foundation models for EEG by systematically distinguishing between learned and engineered electrographic features. Our harmonization protocol involves preparing all recordings across three standard clinical conditions: spontaneous activity, hyperventilation, and photic stimulation. For each condition, we generate multiple data versions to facilitate different analytical strategies. A critical step in our workflow is the transformation of all EEG data into a standard anatomical and functional space, which improves comparability across patients and recordings. Furthermore, to maximize the clinical utility of our dataset, we attach all available clinically relevant labels, including detailed neurological evaluations extracted from EEG reports and formal diagnostic codes.

### Conclusion

Our study presents a research protocol to generate a population-based dataset based on electronic health records in public hospitals, focusing on patients undergoing neurological evaluations. The dataset is centred around routine clinical EEG tests, complemented by neurological reports and diagnostic details, with the potential to incorporate pharmacological data. A key feature of our approach is data harmonization, which standardizes EEG recordings across different stages of routine testing, including photic stimulation and hyperventilation. We also align EEG signals with common functional and anatomical landmarks, ensuring interoperability with other neuroimaging and clinical data sources. These large-scale, standardized datasets provide a foundation for developing AI models and conducting population-level research in clinical neuroscience. They enable new insights into learned representations of EEG activity, facilitate the construction of clinical profiles for neurological patients, and contribute to the development of brain chart reference models of electrophysiological parameters across the lifespan. By enhancing the accessibility and consistency of clinical EEG data, this work supports the broader goal of advancing data-driven approaches in neuroscience and neurology.

## Data Availability

Derivatives of data produced in the present study are available upon reasonable request to the authors

## Institutional Review Board Statement

Approval of all ethical and experimental procedures and protocols was granted by the Research Ethics Board at Simon Fraser University and Fraser Health Authority under Application No. H18-02728.

## Acknowledgments

This research was enabled in part by support provided by the Digital Research Alliance of Canada (alliancecan.ca).

